# Racial Composition in K-12 Schooling and Cognitive Health of Older Black Adults

**DOI:** 10.1101/2024.11.21.24317742

**Authors:** Merci N. Best, Sarah E. Laurent, Cassidy Doyle, Elisa R. Torres, Elham Mahmoudi, Laura B. Zahodne

## Abstract

Systemic educational factors have implications for cognitive health across the life course. Due to *de jure* and *de factor* racial segregation in the United States (U.S.), K-12 school quality varies by racial composition, which we hypothesize contributes to higher risk of Alzheimer’s disease and related dementias among non-Hispanic Black (NHB) older adults. This study adopts a life course perspective to investigate the association between K-12 schools’ racial composition and later-in-life cognitive health among U.S.– born and educated NHB older adults in the Health and Retirement Study (*N*=2,105). Using the 2015, 2017, and 2019 Life History Mail Survey, we categorized racial composition of K-12 schools as only majority non-Hispanic White (NHW) schools (*n*=193), only majority NHB schools (*n*=1,387), or both school types (*n*=525). Multiple linear regressions assessed relationships between school racial composition and total cognition, episodic memory, and vocabulary, controlling for age, gender, childhood factors, and U.S. region of residence at age 10. Attending only NHW schools or both school types was associated with better total cognition (*p*<.01 and *p*<.001, respectively) and episodic memory (*p*<.05 and *p*<.01, respectively) than attending only NHB schools. Attending both school types was also associated with better vocabulary (*p*<.0001). Structural equation models revealed that 45-66% of these associations were mediated by participants’ educational attainment. Our findings highlight the long-term cognitive benefits of attending some NHW schools and underscore the need for initiatives promoting equity in NHB schools. Further, our findings suggest bicultural education may provide language benefits, despite potential interpersonal discrimination toward NHB students.

## Introduction

Lower educational attainment is associated with worse cognitive health in later life, including increased risk for Alzheimer’s disease and related dementia (ADRD).^1^ Non-Hispanic Black (NHB) older adults are twice as likely to develop ADRD compared to their Non-Hispanic White (NHW) counterparts.^1–7^ Racial inequities in educational attainment only partially explain NHB-NHW differences in later-life cognitive health.^8,9^ This finding suggest to the need for better characterization of early educational experiences.^5^ Indeed, well-documented racial inequities in educational quality contribute to NHB-NHW cognitive health disparities above and beyond educational attainment.^10^ Due to structural racism (e.g., historical *de jure* and contemporary *de facto* racial segregation), school racial composition can be a proxy for educational quality. However, it may also reflect exposure to interpersonal discrimination among NHB individuals. Findings regarding associations between school racial composition and later-life cognitive health have been mixed and factors mediating those associations remain unclear.^2,11^

### Segregation influences K-12 school’s racial composition

The 1830s Common School Movement led the way for children to be educated.^12^ Over time, additional reforms including the 1954 U.S. Supreme Court case Brown v Board of Education, began to tackle inequality caused by segregation of students of color.^13^ The 1960s led to Title VI of the Civil Rights Act prohibiting race-based discrimination in schools.^14^ The following year, the Elementary and Secondary Education Act of 1965 provided desegregated school districts with additional federal aid.^15,16^ Desegregating schools through the enforcement of these acts improved NHB student’s academic outcomes by providing them with access to schooling equivalent to their NHW student counterparts.^17^ Seventy years after the Brown v. Board ruling, the percentage of segregation between NHB and NHW students has increased by 64%.^18^ Schools have begun to resegregate for numerous factors, including out-migration of middle-class and affluent NHW people from urban cites to suburbs and the rise of school choice options.^18–20^

Due to structural racism, a school’s racial composition has been considered as a proxy for school quality.^2,17,21–26^ Segregated schools remain separate and unequal, which disproportionately impacts students who attend majority NHB schools that are more likely to have fewer resources, shortage of qualified teachers, larger class sizes, and fewer advanced course options, compared to majority NHW schools.^17,19^ Further, the minimum number of required instruction days in a given school year differs by region; for example, in the U.S. South, NHW schools had 50-100% more instruction days compared to NHB schools until the end of the 1950s.^27^

As a result of the historical state of educational segregation in the U.S., NHB students were and are still more likely to attend NHB schools than NHW students.^17,25,27–29^ While some NHB students attend only NHW schools and have access to higher-quality curricula, they are also more likely to face interpersonal discrimination and structural racism in school environments.^25,29^

### Focus of the current study

Considering the cumulative impact of educational experiences throughout life, we used the life course perspective to examine the association between early educational experiences and cognitive health in later life.^30^ Previous literature on the association between school racial composition and later-life cognitive health has been mixed, but most studies indicate that attending only NHW schools is associated with better cognitive health among NHB older adults.^22,27,31^ However, few studies have focused only on NHB older adults and/or considered attendance of both school types. We hypothesized that NHB older adults who attended only NHW schools would have the highest cognitive health, followed by NHB older adults who attended both school types, and NHB older adults who attended only NHB schools would have the lowest cognitive health. Further, we examined the role of parental educational attainment as a potential mediator of this association. In aim 1, we examine the association between the majority race of K-12 schools attended and cognitive health. Next in aim 2, we examine the moderating role of sex/gender, childhood SES (highest parental education), and U.S. region at age 10. Finally, in aim 3, we examine the mediating role of educational quality (highest educational attainment).

## Materials and Methods

This article does not contain any studies with human or animal participants.

### Analytic sample

The Health and Retirement Study (HRS) is a multistage area probability sample of U.S. adults over age 50.^32^ The HRS is funded by the National Institute on Aging (grant number NIA U01AG009740) and conducted by the University of Michigan.^33^ Primary sampling units were chosen from U.S. Metropolitan Statistical Areas (MSAs) and non-MSA counties. There was over sampling of minorities and the oldest-old. Core interviews were conducted with age-eligible respondents and their spouses approximately every two years since 1992. In addition, the HRS collected information on participants’ schooling history through a Life History Mail Survey (LHMS) in 2015, 2017, and 2019 sponsored by the National Institute on Aging (grant number NIA R01 AG051142) and conducted by the University of Michigan. Details on the sampling strategy are available elsewhere.^34^ The current study was restricted to NHB, U.S.-born adults who reported attending kindergarten through 12^th^ grade, or K-12 schools in the U.S., completed a cognitive assessment in the HRS, and provided data on their school composition during the Life History Mail Survey in 2015, 2017, or 2019 (*N*=2,105). Participants missing responses on the cognitive assessment or school composition were excluded.

### Measures

*Racial composition of school*. Participants were categorized by whether they reported only attending K-12 schools where most of the students were NHW (i.e., a majority NHW K-12 school), only attending K-12 schools where most of the students were NHB (i.e., majority NHB K-12 school), or attending both school types.

Cognitive Health was selected from the wave where they first completed cognitive assessment in the core HRS.

Total cognition was a cognition summary score. Scores range from 0-27 and included immediate and delayed recall of a 10-word list, serial 7s, and backward counting from 20.^35^ Higher scores indicated better cognition.

Episodic Memory was the sum of immediate and delayed free recall of a ten-word list.^35^ For immediate free-recall, the interviewer read a list of 10 nouns (e.g., lake, car, army, etc.), with the participant recalling as many words as possible from the list in any order. For delayed free recall, after approximately 5 minutes of asking other survey questions, the participant was asked to recall the nouns previously presented as part of the immediate free-recall. Scores ranged from 0 to 20, with higher scores indicating better episodic memory.

Vocabulary was used to represent established knowledge, also referred to as crystallized intelligence.^35^ Two different forms with five words each were used from the Wechsler Adult Intelligence Scale-revised (WAIS-R) vocabulary test. Participants were asked to define five words from one of two sets. The interviewer recorded verbatim definitions that were subsequently coded according to instructions for the WAIS-R to indicate the degree of correctness. Responses were coded as 2 if answered perfectly correct, 1 if answered partially correct, and 0 if answered incorrectly. The scores for each of the five items were summed to create a total score ranging from 0 to 10, with higher scores indicating better vocabulary.

*Mediator*. The highest years of education completed by either parent was reported as a continuous variable (range 0-17).

*Covariates*. Age was measured at the time of their first cognitive assessment. Gender was operationalized as a dichotomous variable; female or male. Education was measured by the years of education reported by the participant (range 0-17). Health as a child was dichotomized as excellent/good or fair/poor. The number of books in the childhood home was categorized as none, 1 shelf, 1 or more bookcases, or missing. History of father unemployment during childhood was categorized as yes, no, don’t know, or missing. Region of residence at age 10 was categorized as residing in the South, non-South, or unknown. We examined educational attainment as a mediator and included childhood factors as covariates in line with previous studies, such as educational quality, childhood health, childhood socioeconomic status, and region.^2,11,36,37^

### Statistical Analysis

Descriptive statistics were used to describe the sample mean and standard deviations for continuous variables, number and percent for categorical variables. Differences among participants based on the racial composition of the schools they attended were determined using ANOVA for continuous variables and Chi-square for categorical variables. Separate multiple linear regressions examined associations between the racial composition of K-12 schools attended and the three cognitive outcomes: total cognition, episodic memory, and vocabulary. The same modeling strategy was used for all three outcomes. Model 1 was unadjusted. Model 2 adjusted for age and gender. Model 3 added adjustments for parents’ highest education, region of residence at age 10, number of books in their childhood home, childhood health, and history of father unemployment during childhood. Linear regression modeling was conducted using SAS version 9.4.^38^

Structural equation modeling (SEM) was used to examine the potential mediating associations of educational attainment on the relationship between school racial composition and later life cognitive health. Similar to the modeling strategy described previously, three separate outcomes were examined (total cognition, episodic memory, and vocabulary), and models were adjusted for age and gender. From these models we were able to compute the direct associations of school composition on cognitive health, independent of educational attainment as well as the indirect associations mediated through educational attainment. SEM analyses were conducted using MPlus version 8.2.^39^

## Results

### Sample characteristics

Figure 1 illustrates the schematic flow of our final sample size (*n*=2,105). Table 1 highlights the sample characteristics of NHB older adults who responded to the HRS’s LHMS in 2015, 2017, or 2019 stratified by the racial composition of K-12 schools. About two-thirds of the respondents attended only NHB schools (65.9%), followed by both school types (24.9%). Far fewer participants attended only NHW schools (9.2%). On average, participants were 55 years old when their cognitive health was assessed. Those who attended only NHB schools were on average older (mean= 56) than those who attended only NHW schools (mean= 53, *p*<.001) or both school types (mean= 54, *p*<.001). More years of schooling was completed by those who attended only NHW schools (mean=13.7) or both school types (mean=13.9) on average, than those who attended only NHB schools (mean=12.5, *p*<.001). Those who attended only NHW schools or both school types scored higher on the total cognition assessment (mean=15.8) than those who attended only NHB schools (mean=14.3, *p*<.001).

**Figure 1.**
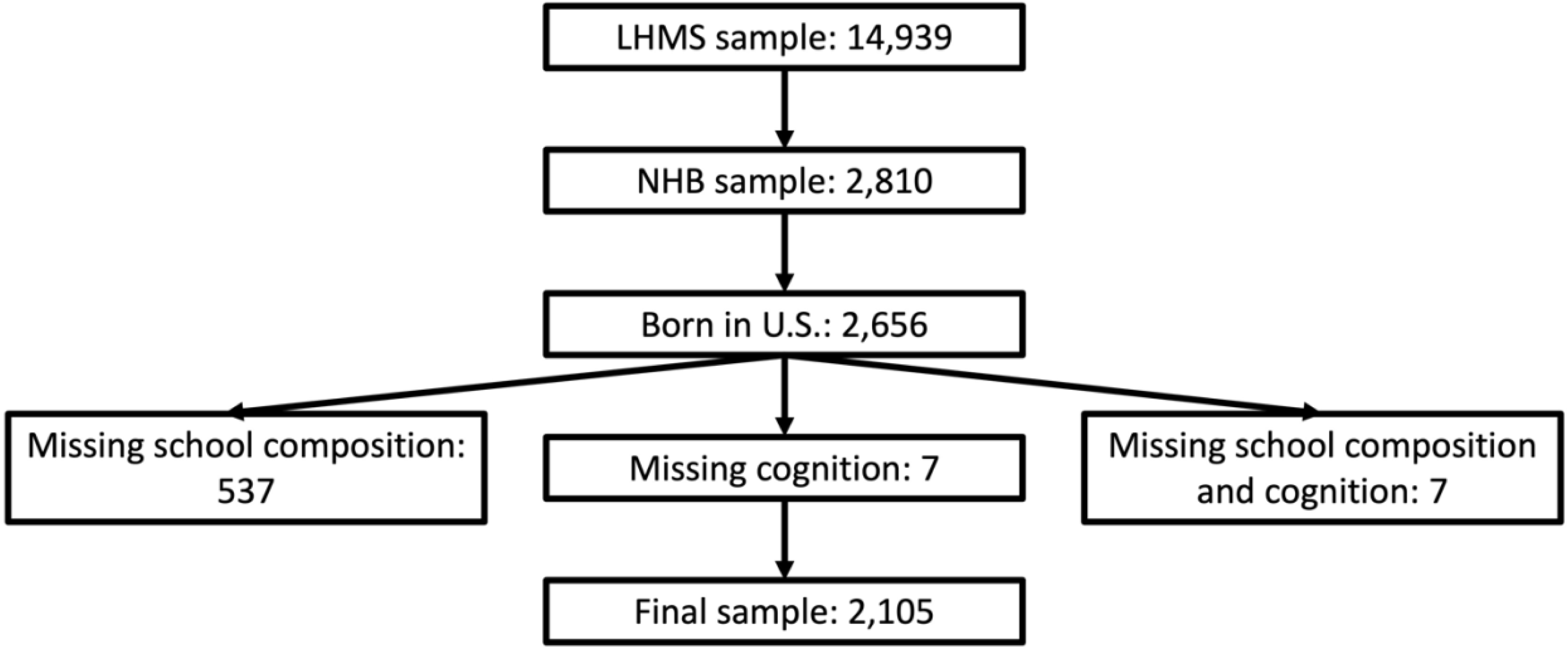
Schematic representation of the derived sample.

**Table 1.** Sample Characteristics.

|  | Only NHW<br>schools<br>(*N* = 193) | Only NHB<br>schools<br>(*N* = 1,387) | Both school<br>types<br>(*N* = 525) | Overall<br>(*N* = 2,105) |  |  |  |
| --- | --- | --- | --- | --- | --- | --- | --- |
|  | M (SD) / %<br>(*N*) | M (SD) / %<br>(*N*) | M (SD) / %<br>(*N*) | M (SD) / % (*N*) | NHW vs.<br>NHB | NHW<br>vs.<br>Both | NHB vs. Both |
| Age<sup>a</sup> | 53.12 (5.5) | 55.74 (6.1) | 53.68 (2.0) | 54.99 (5.9) | <.001 |  | <.001 |
| Gender (F) | 63.73% (123) | 65.61% (910) | 65.90% (346) | 65.51% (1379) | 0.855 | 0.855 | 0.855 |
| Years of education | 13.74 (2.3) | 12.54 (2.6) | 13.89 (2.1) | 12.98 (2.5) | <.001 |  | <.001 |
| Childhood health |  |  |  |  | 0.097 | 0.097 | 0.097 |
| Excellent/Good | 90.16% (174) | 90.63%<br>(1257) | 93.90% (493) | 91.40% (1924) |  |  |  |
| Fair/Poor | 7.77% (15) | 8.07% (112) | 4.57% (24) | 7.17% (151) |  |  |  |
| # of books in house |  |  |  |  | <.001 |  | <.001 |
| None | 13.99% (27) | 40.38% (560) | 18.67% (98) | 32.54% (685) |  |  |  |
| 1 shelf | 22.28% (43) | 23.50% (326) | 22.29% (117) | 23.09% (486) |  |  |  |
| 1+ bookcase | 26.94% (52) | 16.29% (226) | 23.43% (123) | 19.05% (401) |  |  |  |
| Missing | 36.79% (71) | 19.83% (275) | 35.62% (187) | 25.32% (533) |  |  |  |
| Highest parent<br>education | 11.65 (3.1) | 9.85 (3.5) | 11.53 (3.0) | 10.46 (3.4) | <.001 |  | <.001 |
| Father's<br>unemployment |  |  |  |  | 0.052 | 0.052 | 0.052 |
| Yes | 12.44% (24) | 16.01% (222) | 15.62% (82) | 15.58% (328) |  |  |  |
| No | 67.88% (131) | 62.51% (867) | 68.00% (357) | 64.37% (1355) |  |  |  |
| Don't know | 15.54% (30) | 18.82% (261) | 13.14% (69) | 17.10% (360) |  |  |  |
| Missing | 4.15% (8) | 2.67% (37) | 3.24% (17) | 2.95% (62) |  |  |  |
| U.S. region at age 10 |  |  |  |  | <.001 | <.001 | <.001 |
| South | 32.64% (63) | 63.81% (885) | 44.38% (233) | 56.10% (1181) |  |  |  |
| Non-south | 60.10% (116) | 24.73% (343) | 41.71% (219) | 32.21% (678) |  |  |  |
| Unknown | 7.25% (14) | 11.46% (159) | 13.90% (73) | 11.69% (246) |  |  |  |
| *Cognitive health in late life* |  |  |  |  |  |  |  |
| Total cognition<sup>b</sup> | 15.84 (3.7) | 14.34 (4.1) | 15.78 (3.5) | 14.84 (4.0) | <.001 |  | <.001 |
| Episodic memory<sup>c</sup> | 10.50 (2.8) | 9.60 (3.1) | 10.40 (2.9) | 9.88 (3.1) | <.001 |  | <.001 |
| Vocabulary<sup>d</sup> | 5.19 (2.0) | 4.60 (2.1) | 5.46 (2.0) | 4.87 (2.1) | <.001 |  | <.001 |
| <sup>a</sup>Age indicates the age of participants when cognitive health was assessed |  |  |  |  |  |  |  |
| <sup>b</sup>Total cognition is a summary measure including immediate and delayed word recall, backward counting, and serial 7s. Scores range from 0-27 |  |  |  |  |  |  |  |
| <sup>c</sup>Sum of immediate and delayed word recall. Scores range from 0-20. |  |  |  |  |  |  |  |
| <sup>d</sup>Derived from asking participants to define 5 words. Scores range from 0-10 |  |  |  |  |  |  |  |

Similarly, those who attended only NHW schools or both school types scored higher on the episodic memory assessment (mean=10.5 and 10.4, respectively) than those who attended only NHB schools (mean=9.6, *p*<.001). Finally, those who attended only NHW schools or both school types scored higher on the vocabulary assessment (mean=5.2 and 5.5, respectively), than those who attended only NHB schools (mean=4.6, *p*<.001). Other sample characteristics can be found in Table 1.

### Results for School Composition and Total Cognition – Table 2

In the unadjusted model 1, compared to those who attended only NHB schools, participants who attended only NHW schools or both NHW and NHB schools had higher total cognition scores on average (*p*<.001). After adjusting for age and gender in model 2, compared to those who attended only majority NHB schools, participants who attended only NHW schools or both school types still had higher total cognition scores on average (*p*<.001). Model 3 represents the results of our fully adjusted model.

**Table 2.** School Composition and Total Cognition.

|  | Model 1<br>(unadjusted) | Model 2<br>(minimally adjusted) | Model 3<br>(fully adjusted) |
| --- | --- | --- | --- |
| Variable | Estimate (95% CI) | Estimate (95% CI) | Estimate (95% CI) |
| School composition |  |  |  |
| Only NHB schools | - | - | - |
| Only NHW schools | 1.51 (0.91-2.10)*** | 1.43 (0.82-2.01)*** | 1.09 (0.46-1.72)** |
| Both school types | 1.45 (1.05-1.84)*** | 1.36 (0.97-1.76)*** | 1.01 (0.59-1.43)*** |
|  | - | -0.04 (-0.07- -0.01)** | -0.03 (-0.06-0.01) |
| Age |  |  |  |
| Gender |  |  |  |
| Female | - | - | - |
| Male | - | -0.68 (-1.03- -0.32)*** | -0.80 (-1.17- -0.43)*** |
| Highest parent education | - | - | 0.15 (0.09-0.20)*** |
| U.S. region at age 10 |  |  |  |
| South | - | - | - |
| Non-south | - | - | 0.11 (-0.29-0.51) |
| Unknown | - | - | -0.89 (-1.45- -0.32)** |
| Books in household |  |  |  |
| None | - | - | - |
| 1 shelf | - | - | 0.19 (-0.30-0.68) |
| 1+ bookcase | - | - | 0.36 (-0.17-0.89) |
| Missing | - | - | -0.28 (-0.78-0.22) |
| Childhood health |  |  |  |
| Excellent | - | - | - |
| Fair/Poor | - | - | -0.91 (-1.60- -0.23)** |
| Father's unemployment |  |  |  |
| Yes | - | - | - |
| No | - | - | 0.03 (-0.46-0.52) |
| Don't know | - | - | -0.27 (-0.88-0.34) |
| Missing | - | - | 0.46 (-1.01-1.93) |

\*\*\*p<.001; \*\*p<.01; \*p<.05

Compared to participants who attended only NHB schools, those who attended only NHW schools or both school types had higher total cognition scores on average (*p*<.01 and *p*<.001, respectively).

### Results for School Composition and Episodic Memory – *Table 3*

In the unadjusted model 1, compared to those who attended only NHB schools, participants who attended only NHW schools or both NHW and NHB schools had higher episodic memory scores on average (*p*<.001). After adjusting for age and gender in model 2, compared to those who attended only majority NHB schools, participants who attended only NHW schools or both school types still had higher episodic memory scores on average (*p*<.001). In the fully adjusted model, compared to participants who attended only NHB schools, those who attended only NHW schools or both school types had higher episodic memory scores on average (*p*<.05 and *p*<.01, respectively).

**Table 3.** School Composition and Episodic Memory.

|  | Model 1<br>(unadjusted) | Model 2<br>(minimally adjusted) | Model 3<br>(fully adjusted) |
| --- | --- | --- | --- |
| Variable | Estimate (95% CI) | Estimate (95% CI) | Estimate (95% CI) |
| School composition |  |  |  |
| Only NHB schools | - | - | - |
| Only NHW schools | 0.90 (0.44-1.36)*** | 0.08 (0.35-1.26)*** | 0.56 (0.07- 1.04)* |
| Both school types | 0.80 (0.50-1.11)*** | 0.71 (0.41-1.02)*** | 0.49 (0.16-0.82)** |
| Age | - | -0.04 (-0.07- -0.02)*** | -0.03 (-0.06- -0.01)** |
| Gender |  |  |  |
| Female | - | - | - |
| Male | - | -0.82 (-1.09- -0.54)*** | -0.86 (-1.14- -0.57)*** |
| Highest parent education | - | - | 0.08 (0.04-0.13)*** |
| U.S. region at age 10 |  |  |  |
| South | - | - | - |
| Non-south | - | - | 0.13 (-0.18-0.44) |
| Unknown | - | - | -0.45 (-0.89- -0.01)* |
| Books in household |  |  |  |
| None | - | - | - |
| 1 shelf | - | - | 0.32 (-0.06-0.70) |
| 1+ bookcase | - | - | 0.63 (0.22-1.04)** |
| Missing | - | - | -0.10 (-0.48-0.29) |
| Childhood health |  |  |  |
| Excellent | - | - | - |
| Fair/Poor | - | - | -0.95 (-1.48- -0.41)** |
| Father's unemployment |  |  |  |
| Yes | - | - | - |
| No | - | - | -0.08 (-0.46-0.29) |
| Don't know | - | - | -0.15 (-0.62-0.32) |
| Missing | - | - | -0.43 (-1.57-0.71) |

\*\*\*p<.001; \*\*p<.01; \*p<.05

### Results for School Composition and Vocabulary – *Table 4*

In the unadjusted model, compared to those who attended only NHB schools, participants who attended only NHW schools or both NHW and NHB schools had higher vocabulary scores on average (*p*<.001). The scores remain significant and nearly identical in magnitude after adjusting for age and gender in model 2. In the fully adjusted model, compared to participants who attended only NHB schools, those who attended both school types had higher vocabulary scores (*p*<.001). We did not observe a significant association among those who attended majority NHW schools.

**Table 4.** School Composition and Vocabulary.

|  | Model 1<br>(unadjusted) | Model 2<br>(minimally adjusted) | Model 3<br>(fully adjusted) |
| --- | --- | --- | --- |
| Variable | Estimate (95% CI) | Estimate (95% CI) | Estimate (95% CI) |
| School composition |  |  |  |
| Only NHB schools | - | - | - |
| Only NHW schools | 0.59 (0.27-0.92)*** | 0.61 (0.29-0.94)*** | 0.20 (-0.14-0.54) |
| Both school types | 0.86 (0.65-1.08)*** | 0.88 (0.66-1.10)*** | 0.60 (0.37-0.83)*** |
| Age | - | 0.01 (-0.01-0.02) | 0.02 (0.00-0.04)* |
| Gender |  |  |  |
| Female | - | - | - |
| Male | - | 0.02 (-0.18-0.21) | 0.01 (-0.19-0.21) |
|  | - | - | 0.01 (0.06-0.13)*** |
| Highest parent education |  |  |  |
| U.S. region at age 10 |  |  |  |
| South | - | - | - |
| Non-south | - | - | 0.42 (0.21-0.64)*** |
| Unknown | - | - | -0.08 (-0.38-0.23) |
| Books in household |  |  |  |
| None | - | - | - |
| 1 shelf | - | - | 0.16 (-0.11-0.43) |
| 1+ bookcase | - | - | 0.57 (0.28-0.86)*** |
| Missing | - | - | -0.02 (-0.29-0.25) |
| Childhood health |  |  |  |
| Excellent | - | - | - |
| Fair/Poor | - | - | -0.42 (-0.79- -0.05)* |
| Father's unemployment |  |  |  |
| Yes | - | - | - |
| No | - | - | -0.07 (-0.34-0.19) |
| Don't know | - | - | 0.16 (-0.18-0.49) |
| Missing | - | - | 0.27 (-0.54-1.07) |

\*\*\*p<.001; \*\*p<.01; \*p<.05

Figure 2 shows the results from SEM. The figures display the standardized estimates examining the mediating associations of educational attainment on the association between school racial composition and the three cognitive outcomes (total cognition, episodic memory, and vocabulary). In, panel A, the outcome is total cognition. We observed an indirect association of attending only majority NHW schools on total cognition through educational attainment (*p*<0.001), with 45.2% of the total associations mediated. A direct association of attending only majority NHW schools on total cognition independent of educational attainment remained (*p*=0.006). Similarly, we observed an indirect association of attending both school types on total cognition through educational attainment (*p*<0.001), with 53.3% of the total associations mediated. A direct association of attending both school types independent of educational attainment remained (*p*=0.001).

**Figure 2.**
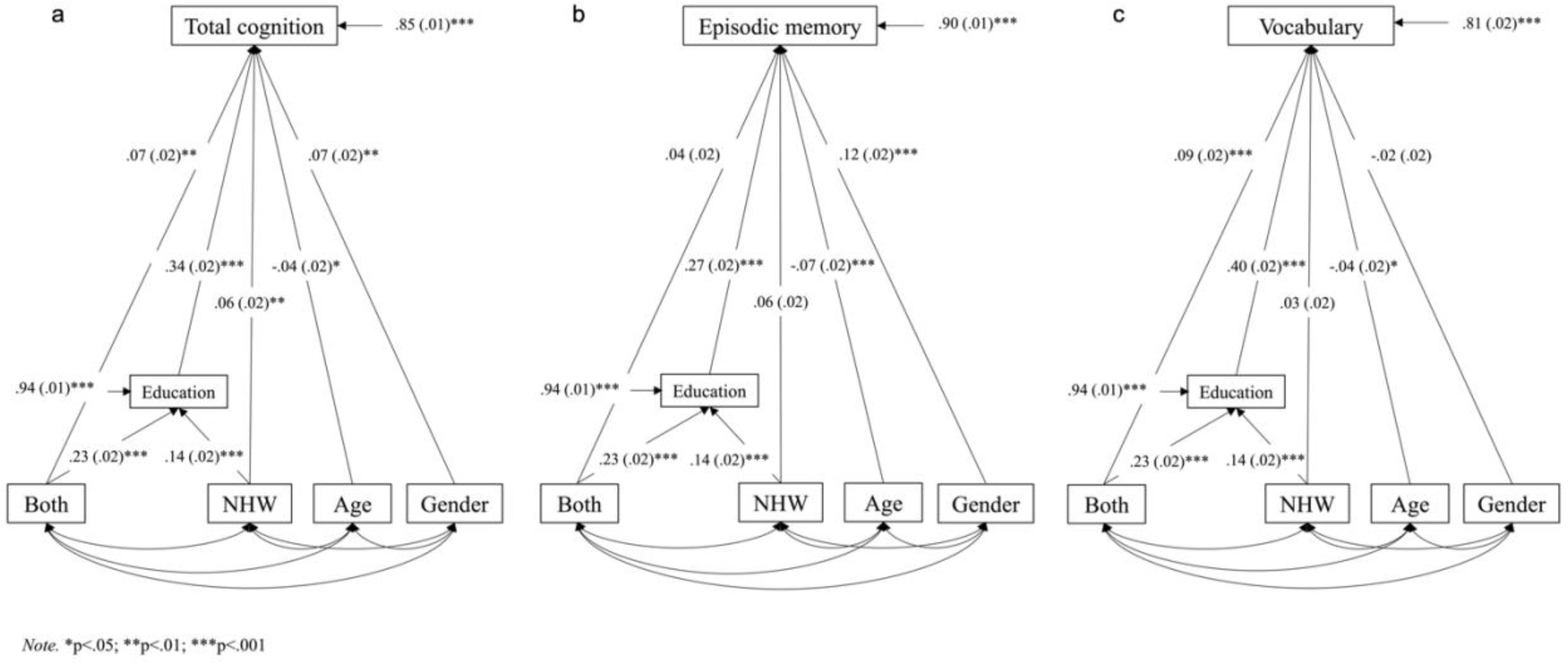
Structural equation modeling of the associations between school’s racial composition and cognitive health. A) Total cognition. B) Episodic memory. C) Vocabulary.

In panel B, the outcome is episodic memory. We observed an indirect association of attending only majority NHW schools on episodic memory through educational attainment (*p*<0.001), with 48.1% of the total associations mediated. Similarly, we observed an indirect association of attending both school types on episodic memory through educational attainment (*p*<0.001), with 61.8% of the total associations mediated.

Finally, in panel C the outcome is vocabulary. We observed an indirect association of attending only majority NHW schools on vocabulary through educational attainment (*p*<0.001), with 65.9% of the total association mediated. Similarly, we observed an indirect association of attending both school types on vocabulary through educational attainment (*p*<0.001), with 51.9% of the total association mediated. A direct association of attending both school types independent of educational attainment remained (*p*<0.001).

## Discussion

Our within-group analysis provides additional evidence of the positive association between attending racially discordant schools and cognitive health outcomes in NHB older adults. These findings advance the literature by showing better cognitive health among NHB older adults attending some or only majority NHW schools, while also demonstrating a potential benefit of bicultural education to vocabulary.

In aim 1, we examine the association between the majority race of K-12 schools attended and cognitive health. Consistent with prior literature using community samples or the HRS-LHMS dataset, we provide additional evidence that attending some or only majority NHW schools is associated with higher total cognition scores.^11,22,27^ Additionally, attending some or only majority NHW schools is associated with higher episodic memory scores.^11,25,27^ Our study advances the literature by including a third K-12 school racial composition group that attended both majority NHW and majority NHB schools. Individuals in a both school types group likely have complex schooling experiences. While these students benefit from higher quality education when attending majority NHW schools, they also experience less discrimination when attending majority NHB schools. By including a both school types group, our work provides a more comprehensive understanding of NHB older adults that can inform educational policies on school integration and diversity. Our results suggest that the beneficial effects of higher school quality outweigh the possible detrimental effects related to NHB students being exposed to discrimination while attending only majority NHW schools. We hypothesize that these students also benefit from an ability to code-switch or alternate between the use of Standard American English, a formal form of English with fixed norms for spelling, grammar, and usage that is taught in schools, and African American Vernacular English, a linguistic structure with different grammatical, phonological, and lexical features from Standard American English.^40–42^

We also advanced the literature by introducing vocabulary scores as a third cognitive health outcome to help capture the linguistic and verbal aspects of cognition that may not be fully assessed by total cognition and episodic memory scores. Further, only attending some majority NHW schools is associated with higher vocabulary scores. We hypothesize that these vocabulary findings are linked to previous studies indicating that being regularly exposed to lexicons from multiple cultures may result in larger vocabulary.^43,44^ NHB children who attend both school types, likely benefit from exposure to lexicons during their time in majority NHW schools and majority NHB schools, resulting in higher vocabulary scores.

In aim 2, we examine the moderating role of sex/gender, childhood SES (highest parental education), and U.S. region of residence at age 10. We found that K-12 school’s racial composition and cognitive health also differed by highest parent education, U.S. region of residence at age 10, and childhood health. Total cognition and memory also differed by gender. Specifically, females have higher total cognition and memory scores than males, which is consistent with some of the literature that suggest that the gendered ways of socializing young children in schools favor girls ^45–47^.

In aim 3, we examine the mediating role of educational quality, specifically highest educational attainment. Higher educational quality is more likely to occur in well-resourced schools that often have larger NHW student populations.^16^ In our study, a significant portion of the associations between school’s racial composition and cognitive health was attributed to educational attainment. Specifically, those who attend some or only majority NHW schools had more years of education on average. In turn, more years of education was associated with better cognitive health.

The identification of educational attainment as a mediator of school composition-cognition associations points to another potential point of intervention to interrupt risk pathways for NHB older adults who attended lower-quality schools. As schools have resegregated in the U.S. since the 1990s, additional legislation has sought to provide benefits to all students.^19^ NHB students can benefit from comprehensive, multifaceted policies that reduce educational segregation and address disparities. For example, special programs to promote high school completion and facilitate college attendance could be targeted to those individuals.

These findings highlight the long-term cognitive benefits of attending some NHW schools while underscoring the need for initiatives promoting equity in NHB schools. Broader improvements in access to higher-quality schooling may reduce educational disparities and manifest as better cognitive health in later life. Given that the current results are perhaps indicative of cognitive decline, we hypothesize that policies aimed at improving early educational experiences may positively influence NHB older adult’s cognitive health later in life. Ensuring a good education for all American students could result in a 7% reduction in ADRD prevalence, which would greatly benefit society.^1^

### Limitations

Our inclusion criteria of the HRS-LHMS sample present both a limitation and a strength. Because the LHMS does not have weights the current study is not representative of the general population. Specifically, we do not know if these findings are generalizable to other racial groups or even NHB individuals of Hispanic ethnicity, not born in the U.S. and/or not educated in the U.S. Importantly, our limited, controlled, and homogenous sample allows us to better isolate the associations between K-12 school’s racial composition and cognitive health. The U.S.-born and educated NHB older adults included in our sample share a more consistent historical experience of segregation and systematic discrimination in the U.S., which makes our insights and recommendations more targeted.

This dataset also does not provide objective metrics of school quality, for example, per-pupil spending or level of local investment in schools. Our results are consistent with studies that have used more objective measures of school racial composition and school quality.^28,31^ Further, while we attempted to examine potential domain-specificity by isolating the episodic memory component of total cognition measure and adding vocabulary scores as an additional outcome, another limitation of our study is the relatively limited cognitive battery.

The HRS does not provide data on cognitive health before age 50, therefore we conducted a cross-sectional analysis. While our study design cannot address how cognitive health changes throughout the life course, other longitudinal studies show that effects are stronger for the level of cognitive health than for rate of change in cognition over time.^48^ Finally, although there may have also been additional uncontrolled confounders, we leveraged the entire HRS-LHMS to incorporate relevant childhood characteristics into our models.

Future studies could benefit from using more comprehensive neuropsychological assessments to provide higher-quality measurement of the constructs included in this study, as well as additional constructs not well represented in the core HRS battery. In these studies, it would also be interesting to further characterize the both school types group by the precise durations and grade levels of attendance at majority NHW versus majority NHB schools. Additionally, it would be interesting to compare the current cohort to future generations to assess whether segregation today has the same effects on NHB students. These future studies would also benefit from incorporating a longitudinal design to assess the longer-term impact of K-12 schools’ racial composition on cognitive health. Finally, within these studies, it would also be interesting to incorporate an intervention to test its efficacy in improving students attending only majority NHB schools’ cognitive health outcomes.

In conclusion, this study provides evidence of the benefits of attending some or only NHW schools earlier in life on cognitive health later in life among U.S.-born and educated NHB older adults. Our study’s findings further emphasize how bicultural education can uniquely serve as a benefit to NHB older adult’s vocabulary scores across the life course. Considering less education is an early modifiable risk factor for ADRD it is imperative to address how inequities in K-12 school experience increase ADRD risk in later life, especially for NHB older adults who are twice as likely to be at risk for ADRD compared to NHW older adults.^1^ NHB students will likely benefit from the growing efforts in the U.S. to ensure all students have equitable access to a high-quality education, regardless of their school’s racial composition.

## Ethics and reporting

The authors declare that all relevant ethical guidelines have been followed, all necessary IRB and/or ethics committee approvals have been obtained, all necessary patient/participant consent has been obtained and the appropriate institutional forms archived.

## Competing interest statement

None of the authors or their institutions received any payments or services in the past 36 months from a third party that could be perceived to influence, or give the appearance of potentially influencing, the submitted work.

## Funding statement

The authors disclosed receipt of the following financial support for the research, authorship, and/or publication of this article: this work was supported by the National Institutes of Health T32 NS007222 (M.N.B). Funding for this research was also made possible in part by the Michigan Pioneers Fellows Program at the University of Michigan (M.N.B). This work was also supported by two grants from the Alzheimer’s Association 24AARFD-1189285 (M.N.B) and AARGD-NTF-21-848187 (E.R.T.). This work received support from the Health and Retirement Study (HRS) grant, and the HRS affiliated R01 (AG-051142) that funded the development of the Life History Mail Survey, coding, and dissemination of the 2015-2017 cross-wave file together with the 2019 wave data.

Research reported in this publication was supported by the National Institutes of Health through the Michigan Center for the Contextual Factors in Alzheimer’s Disease (P30AG059300) and the HRS (U01AG009740). The content is solely the responsibility of the authors and does not necessarily represent the official views of the National Institutes of Health. The HRS is sponsored by the National Institute on Aging (NIA; U01AG009740) and is conducted by the University of Michigan. HRS, public use dataset. Produced and distributed by the University of Michigan with funding from the National Institute on Aging (grant number NIA U01AG009740). Ann Arbor, MI, (2024). None of the authors or their institutions at any time received payment or services from a third party for any aspect of the submitted work.

## Author Contributions

Merci Best (Conceptualization; Project Administration; Writing – Original Draft; Writing – Review and Editing); Sarah Laurent (Conceptualization; Data Curation; Formal Analysis; Writing – Review and Editing); Cassidy Doyle (Conceptualization; Validation; Writing – Review and Editing); Elisa Torres (Conceptualization; Writing – Original Draft; Writing – Review and Editing); Elham Mahmoudi (Conceptualization; Writing – Review and Editing); Laura Zahodne (Conceptualization; Writing – Review and Editing).

## Data Availability

All data produced in the present study are available upon reasonable request to the authors.

https://hrs.isr.umich.edu/about

## Acknowledgements

The authors would like to thank the 2024 Michigan Center for Contextual Factors in Alzheimer’s Disease (MCCFAD) Summer Data Immersion Program directors, Kristine Ajrouch, Toni Antonucci, and Laura Zahodne; Research and Education Core co-leads, Sheria Robinson-Lane and Hwajung Choi; MCCFAD Analytic Core co-leads, Richard Gonzalez and Noah Webster; and Survey Research Center for the Institute for Social Research (ISR) Research scientist Amanda Sonnega for the analytic training, exposure to secondary data sources, networking opportunities, and technical expertise. We also thank all the Health and Retirement Study (HRS) panel participants, ISR Survey Research Operations, HRS production staff, and the life history mail survey research team.

## Datasets/Data Availability

The data supporting the findings of this study are available on request from the corresponding author.

## Notes

### Competing Interest Statement

The authors have declared no competing interest.

### Author Declarations

The study used ONLY openly available human data that were originally located at: https://hrs.isr.umich.edu/about

